# Suicide Risk in Borderline Personality Disorder: a Machine Learning Tool based on Clinical and MRI Data

**DOI:** 10.1101/2024.07.25.24310985

**Authors:** Claudio Crema, Alberto Boccali, Alessandra Martinelli, Silvia De Francesco, Serena Meloni, Cesare M. Baronio, Laura Pedrini, Mariangela Lanfredi, Damiano Archetti, Alberto Redolfi, Roberta Rossi

## Abstract

Borderline Personality Disorder (BPD) is a complex mental condition. Individuals with BPD have an average of three lifetime suicide attempts, and 10% of them die by suicide. Understanding risk factors linked to suicidal behaviors is crucial for effective intervention strategies. In recent years, machine learning (ML) approaches for predicting suicide risk in persons with mental disorders have been developed, but a reliable, BPD-specific tool is lacking. In this work, we developed DRAMA-BPD (Detecting Risk factors for suicide Attempts with Machine learning Approaches in Borderline Personality Disorder), a second-opinion tool to assess suicide risk in individuals with BPD. DRAMA-BPD, built upon a Support Vector Machine (SVM) classifier, is trained on the CLIMAMITHE (CLM) dataset, which encompasses sociodemographic, clinical, emotional assessments, and MRI data. Feature selection revealed that 6 out of the 7 most important features are MRI-derived, and a comprehensive review was conducted to ensure consistency with existing scientific literature. The classifier achieved an overall Area Under the Curve (AUC) of 0.73, Precision (P) of 0.75, Recall (R) of 0.70, and F1-score of 0.72. Tests were conducted on the independent SUDMEX_CONN dataset, yielding an AUC of 0.59, P of 0.46, R of 0.92, and F1 of 0.62. While there is a significant imbalance between Precision and Recall, these results demonstrate the potential utility of the proposed model.

## 1. Introduction

Borderline Personality Disorder (BPD) is a severe illness characterized by emotional instability, fear of abandonment, chronic emptiness, identity disturbance, impulsivity, intense anger, unstable relationships, self-harm, and transient paranoid ideation or severe dissociative symptoms [1]. BPD individuals typically exhibit an average of three lifetime suicide attempts than general population, predominantly involving overdose [2-5], and research indicates that up to 10% of them die by suicide. Notably, emotional dysregulation, a hallmark characteristic of BPD, plays a substantial role in augmenting the susceptibility to suicidal behaviors within this clinical population [6-8]. Conversely, the self-harm, such as cutting, represents a typical manifestation of non-suicidal self-injury aimed at temporary emotional relief or reducing dissociative states, rather than indicating a desire for death. Understanding the specific criteria and risk factors linked to suicide attempts in individuals with BPD is crucial for effective suicide risk assessment and intervention strategies [9].

In recent years, the use of Machine Learning (ML) algorithms for identifying and predicting suicide risk has been rapidly expanding. These algorithms can be trained to classify and forecast future suicide attempts by utilizing data from diverse sources and integrating a wide array of clinical, neurobiological, behavioral, and social risk factors. In a recent systematic review [10], 81 studies using ML techniques to assess suicide risk or predict suicide attempts in individuals with mental disorders were examined. Despite methodological differences, most studies reported accuracies of 0.7 or higher, considering factors like previous attempts, disorder severity, and pharmacological treatments; however, only 3 of these studies tested their prediction on independent datasets. Only a limited number of studies have employed ML specifically to address suicide risk on the BPD population, and just a few of them use Magnetic Resonance Imaging (MRI) data. One is the study by Tian et al. [11], where a k-NN model was developed on resting-state functional MRI variables of individuals with bipolar disorder (overall population composed by 288 subjects), resulting in a final suicide prediction accuracy of 0.72 on an independent dataset. From a ML methodology point of view, many of the previously mentioned works present some limitations. Some may be overfitting the training set due to an excessive number of input features. A model tends to overfit the training dataset when dealing with too many features; this phenomenon, known as the “curse of dimensionality,” undermines their general applicability [12]. Another limitation in the literature is the lack of validation on external independent datasets [13]. External validation is an important aspect to consider when developing a ML model, as it estimates performance on populations different from the training one, allowing to make assessments on the generalizability of the model [14]. It is also an effective way to check if the model is overfitting.

To address the literature gaps, this study aims to develop “Detecting Risk factors for suicide Attempts with Machine learning Approaches in Borderline Personality Disorder” (DRAMA-BPD), a ML-based tool for suicide prediction in individuals with BPD by integrating clinical standardized assessments with MRI data, i.e. T1-weighted three-dimensional (T13D), Diffusion-Weighted Imaging (DWI), and Fluid-Attenuated Inversion Recovery (FLAIR). Every step of the ML pipeline implementation has been carefully analyzed, in order to follow methodological guidelines. The findings of this research have the potential to facilitate the development of models suitable for clinical implementation.

### 2. Methods

The study design comprised several stages: initial data preprocessing, identification of significant features, subject classification. MRI data underwent automated processing to extract various brain region characteristics such as volumes, cortical and subcortical thicknesses, WM lesions, and WM diffusion metrics. The values extracted were then utilized, alongside with other data, to train and evaluate the DRAMA-BPD model, employing a Leave-One-Out (LOO) strategy for classification into Suicide Attempters (SAs) and Non-Attempters (NAs). Finally, in order to test the general applicability of DRAMA-BPD, we tested it on an independent dataset.

### 2.1 Data

The total number of subjects in the dataset is 60, balanced between SAs (30 subjects, the exact 50% of the dataset) and NAs. The total number of features is 276. They can be divided in 3 separate groups:

- Sociodemographic and Proxy Clinical (SPC) features, composed of sociodemographic data (e.g., sex, marital status), and indicators of clinical information (e.g., onset age, alcohol/substance abuse). The total number of SPC features is 19;
- Clinical features, composed by results of several tests, described in paragraph 2.2.1. The total number of Clinical features is 48;
- MRI features, an aggregation of volumes and thicknesses of subcortical regions, along with MDs, FAs, and data of lesions provided by LPA. The total number of MRI features is 209.

#### 2.1.1 Clinical and Emotional Assessments

The data utilized in this study were sourced from an investigation on individuals with BPD, i.e., the CLIMAMITHE Study (CLM, [15, 16]), a longitudinal, multicenter, randomized clinical trial (Clinical Trials.org as NCT02370316) conducted at IRCCS Istituto Centro San Giovanni di Dio, Brescia, Italy (coordinating center) and the Third Center of Cognitive Psychotherapy, Rome, Italy - Scuola Italiana di Cognitivismo Clinico (SICC), Rome; the study evaluates the clinical and neurobiological impact of Metacognitive Interpersonal Therapy (MIT) on individuals with BPD. The protocol was approved with number 67/2014. CLM included subjects i) who met the DSM-5-TR criteria required for a BPD diagnosis, ii) aged between 18 and 45 years and iii) able to speak and write in Italian. Individuals excluded were those i) who refused to provide informed consent, ii) with a lifetime diagnosis of schizophrenia, schizoaffective disorder, substance abuse or dependence within the three months prior to enrollment, bipolar disorder, organic mental syndromes, dementia, or cognitive impairment, along with relevant neurological symptoms, iii) who were pregnant or lactating, iv) who were currently undergoing psychotherapy. Trained psychologists conducted a multidimensional assessment covering clinical and emotional aspects, using several standardized tools. Structured Clinical Interview for DSM (SCID II) [17], Zanarini Rating Scale for Borderline Personality Disorder (ZAN-BPD) [18], and Difficulties in Emotion Regulation Scale (DERS) [19] [20] are some of the tools utilized. Furthermore, Data on sociodemographics, family psychiatric and trauma history, suicide attempts, self-harm and aggression episodes, hospitalizations, alcohol and substance abuse, medical comorbidities, pharmacotherapy, and psychotherapy interventions were collected.

#### 2.1.2 Magnetic Resonance Imaging

The CLM database contains T13D, DTI, and FLAIR images from a Siemens scanner (Skyra, 3T) obtained at the Spedali Civili of Brescia^1^. The image processing pipelines employed included FreeSurfer (FS-recon-all), version 7.3.2, TRActs Constrained by UnderLying Anatomy (TRACULA), and Lesion Prediction Algorithm (LPA). FS facilitates the segmentation of cortical and subcortical brain structures from T13D images, which were corrected for smooth intensity variations using N4BiasFieldCorrection from Advanced Normalization Tools (ANTs) and pre-registered to the MNI_152_T1_1mm template using the FSL *flirt* function (12 DOF) [21, 22]. The N4 bias field correction algorithm is a popular method for correcting low frequency intensity non-uniformity present in MRI image data [23]. In this study, among all the possible output variables from FS, subcortical volumes and cortical thicknesses were selected as features of interest. Quality control of the output was performed retrospectively, inspecting the results of each pipeline slice by slice and discarding those of low quality or incorrect segmentation.

### 2.2 Classifier pipeline

Given the dataset containing 276 features and 60 subjects, this would result in extreme overfitting; therefore, the first step was feature selection, with the aim to reduce the number of features to 7. First, we removed features with more than 50% missing values. We then split the dataset into training and test set, with the test set being 30% of the total dataset. We performed encoding of categorical features, scaling with MinMaxScaler from Python scikit-learn library [24] and data imputation of missing values by means of a k-Nearest Neighbors imputer. We executed feature selection by first removing heavily correlated features using the Variance Inflation Factor (VIF) with a threshold of 10. Subsequently, we selected the most important features with a RF classifier with 100 estimators. After defining the most important features, we implemented a SVM pipeline by following a standard approach, i.e. a Leave-One-Out (LOO) classification, performing a Grid Search to find the optimals parameters for each split. Finally, we performed a power analysis to estimate the sample size required to achieve performance levels reported in the literature using a technique called Nx Subsampling [25].

### 2.3 Independent test sets

In order to test the general applicability of DRAMA-BPD, we ran a test on an independent dataset called SUDMEX_CONN [26], MEX from now on, the result of a case-control study of individuals with Cocaine Use Disorder (CUD). The dataset is composed of 145 subjects (64 HC, 74 CUD, and 7 with unknown status), with a large number of neuropsychiatric tests, e.g., DERS, SCID. Additionally, MRI sequences have been taken for these subjects: T1-weighted, 10 minutes rsfMRI, and HARDI-DWI multishell, acquired with a Philips scanner (Ingenia, 3T) obtained at the National Institute of Psychiatry in Mexico City. See Table 1 for more information. Due to the low quality of the original DTI images, for proper registration to the MNI template, it was necessary, following N4 correction and eddy currents removal, to introduce, before applying the *flirt* function, a manual 9 DOF registration of the DTI to the JHU-ICBM-DWI-2mm template. Quality control was performed for MEX similarly to CLM. Finally, the neuroimaging data from the two databases were harmonized using the *harmonizationLearn* function (part of the Python package neuroHarmonize, [27]), with CLM data as reference, in order to remove differences arising from the different types of scanners used during acquisition. To be compatible with DRAMA-BPD, we employed the TLP item in the SCID scale to identify subjects with BPD. The TLP item represents a dual variable indicating the presence or absence of BPD. We then used the C9 item of the Mini-International Neuropsychiatric Interview (MINI) scale to identify SAs [28]. Finally, subjects with available MRI data were selected. This resulted in a total of 32 subjects, 19 of whom NA and 13 SA.

**Table 1.** MRI typologies and features in CLM and MEX datasets.

| Study | Scanner | T13D |  | FLAIR |  | DTI |  |
| --- | --- | --- | --- | --- | --- | --- | --- |
|  |  | Field Strength (T) | Pix Dim (mm) | Field Strength (T) | Pix Dim (mm) | Field Strength (T) | Pix Dim (mm) |
| CLM | Siemens - Skyra | 3.0 | 1x1x1 | 3.0 | 0.9x0.9x4 | 3.0 | 2x2x2 |
| MEX | Philips - Ingenia | 3.0 | 1x1x1 | N/A | N/A | 3.0 | 2x2x2 |

## 3. Results

### 3.1 Classifier pipeline

The most important features resulting from the selection process are the following:

- Thickness of the right medial OrbitoFrontal Cortex (OFC)
- Number of lifetime admissions to SPDC (NumSPDC)
- FA average weight of Anterior Commissure (AC)
- Hypointensities of the WM
- Volume of the right caudate (RC)
- Volume of the corpus callosum posterior (CCP)
- Volume of the left choroid plexus (LCP)

We analyzed the differences of these features between SAs and NAs. For every feature we calculated the average value and standard deviation for SA and NA populations, obtaining then the p-value with Kruskal-Wallis test [29]. Results are reported in Table 2.

**Table 2.** Values of the seven most important features for SA and NA groups. Values in brackets indicate the number of subjects for whom the characteristic is available.

| Feature | SA (30 subjects) | NA (30 subjects) | P-value |
| --- | --- | --- | --- |
| Thickness right medial OFC [mm] | $2.55 \pm 0.15$ (26) | $2.41 \pm 0.12$ (29) | 0.0010 |
| NumSPDC | $1.50 \pm 2.91$ (30) | $0.27 \pm 0.83$ (30) | 0.0024 |
| FA average weight AC | $0.35 \pm 0.02$ (30) | $0.36 \pm 0.02$ (29) | 0.0364 |
| Hypointensities WM [mm<sup>3</sup>] | $1750 \pm 397$ (26) | $1550 \pm 343$ (30) | 0.0224 |
| Volume RC [mm<sup>3</sup>] | $4635 \pm 590$ (27) | $4829 \pm 465$ (30) | 0.2632 |
| Volume CCP [mm<sup>3</sup>] | $1219 \pm 137$ (27) | $1275 \pm 155$ (30) | 0.2843 |
| Volume LCP [mm<sup>3</sup>] | $646 \pm 202$ (27) | $558 \pm 140$ (30) | 0.0531 |

Among the most important features, no Clinical are listed, while one of them is a SPC feature (NumSPDC), and 6 out of 7 are MRI features. Considerations about the retrieved important features will be analyzed in the Discussion section. The confusion matrix of DRAMA-BPD pipeline results is shown in Figure 1. The classification by means of LOO resulted in the following global metrics: accuracy = 0.733, Precision = 0.750, Recall = 0.700, F1-score = 0.724, AUC = 0.733. The number of False Negatives (SA predicted as NA) is slightly larger than the number of False positives (NA predicted as SA), causing the Precision being slightly higher than Recall.

**Figure 1.**
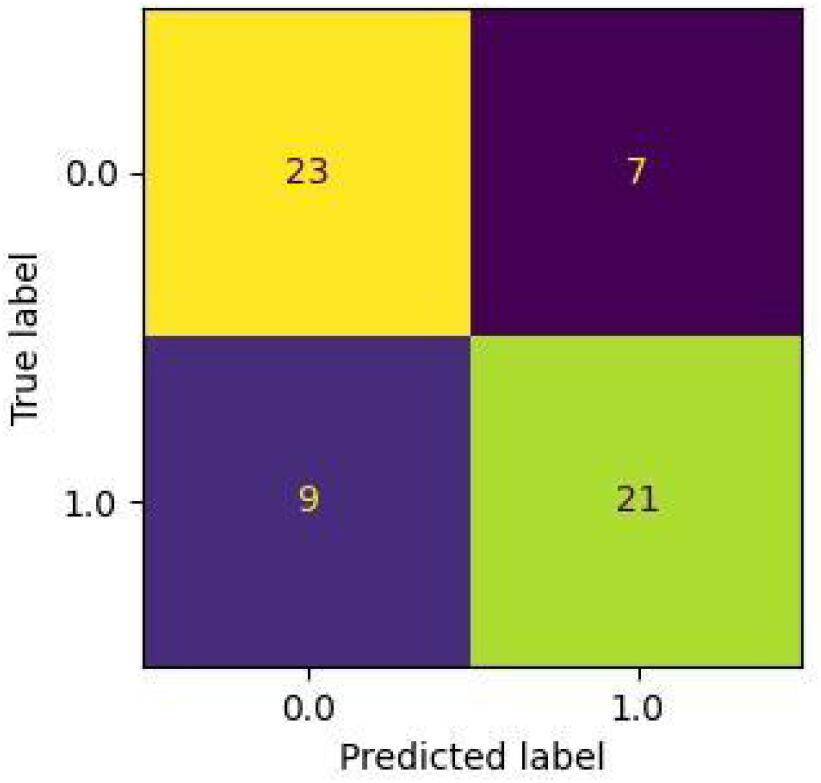
Confusion matrix of the LOO classification on the CLM dataset

The power curve has been calculated for four points of the training set (that corresponds to 75% of the CLM dataset): 25%, 50%, 75% and 100%, corresponding to 11, 22, 33, and 45 records, respectively. To prevent overfit, the number of features has been reduced accordingly; 1, 2, 4, and 7 features, respectively, from most to least important. Results for accuracy are the following:

- 25% of the training set: 0.582 ± 0.067
- 50% of the training set: 0.642 ± 0.102
- 75% of the training set: 0.727 ± 0.108
- 100% of the training set: 0.692 ± 0.116

The power curve is visible in Figure 2. Results show that to reach a target accuracy of 0.85, in line with the most performing literature works, the original dataset size should be increased by four times, reaching a total of approximately 250 records.

**Figure 2.**
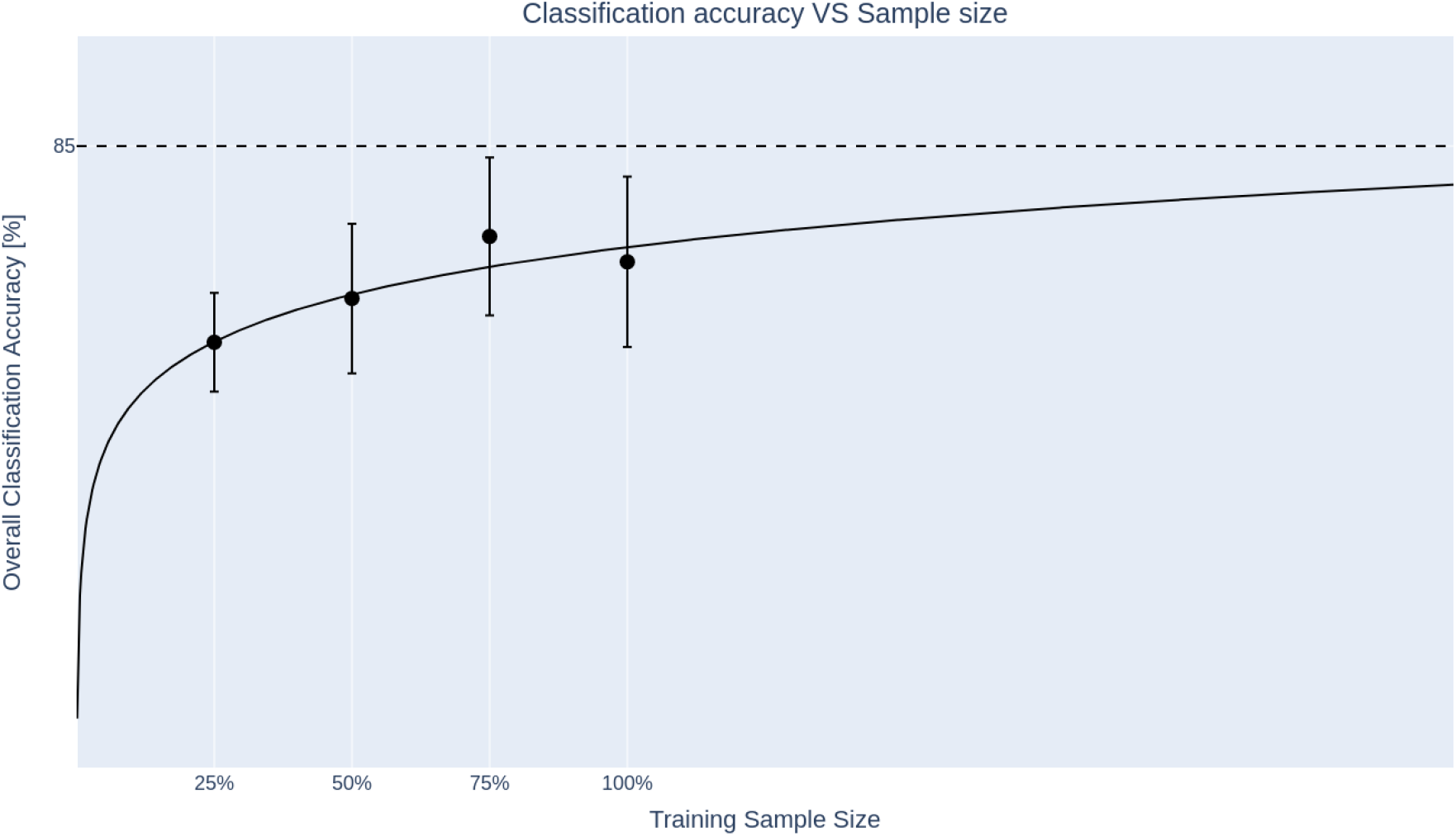
Classifier power curve.

### 3.3 Independent test sets

We conducted statistical analyses to compare the CLM and MEX datasets based on age, sex, and CUD. Intra-dataset comparisons (SA vs NA for both CLM and MEX) are presented in Table 3, while inter-dataset comparisons (CLM SA vs MEX SA and CLM NA vs MEX NA) are shown in Table 4. Intra-dataset tests showed no statistical differences for either of the features. Conversely, inter-dataset tests revealed significant differences in sex and CUD for both SA and NA. Corrections were necessary due to these differences, as discussed in Section 2.3 “Independent test set”. Specifically, we identified the same exact features used by the DRAMA-BPD classifier for comparison. All 6 MRI features are present for the 32 MEX subjects. To address the absence of the “number of lifetime admissions to SPDC” feature in MEX (SPDC do not exist in Mexico), we substituted it with the P1 item of the Addiction Severity Index (ASI) scale [30]. This scale, used in addiction treatment and research, evaluates an individual’s substance use and related problems across seven domains (e.g., drugs, alcohol). The P1 item of this scale asks “*How many times have you been treated for any psychological or emotional problem in a hospital?*”.

**Table 3.** Comparison for age, sex, and CUD features between SA and NA groups for CLM and MEX datasets. Values in brackets indicate the number of subjects for whom the characteristic is available.

| Feature | CLM |  |  | MEX |  |  |
| --- | --- | --- | --- | --- | --- | --- |
|  | SA (30) | NA (30) | p-value | SA (13) | NA (19) | p-value |
| Age (years) | 29.3 ± 8.7<br>(30) | 29.2 ± 6.9<br>(30) | 0.728 | 30.5 ± 6.2<br>(13) | 29.9 ± 7.0<br>(19) | 0.759 |
| Sex (% of females) | 90% (30) | 83.3% (30) | 0.459 | 0% (13) | 15.8% (19) | 0.149 |
| CUD (%) | 26.7% (30) | 23.3% (30) | 0.775 | 100% (13) | 100% (19) | 1.000 |

**Table 4.** Comparison for age, sex, and CUD features between CLM and MEX datasets for SA and NA groups. Values in brackets indicate the number of subjects for whom the characteristic is available.

| Feature | SA |  |  | NA |  |  |
| --- | --- | --- | --- | --- | --- | --- |
|  | CLM (30) | MEX (13) | p-value | CLM (30) | MEX (19) | p-value |
| Age (years) | 29.3 ± 8.<br>(30) | 30.5 ± 6.2<br>(13) | 0.4423 | 29.2 ± 6.9<br>(30) | 29.9 ± 7.0<br>(19) | 0.6660 |
| Sex (% of females) | 90% (30) | 0% (13) | <0.0001 | 83.3% (30) | 15.8% (19) | <0.0001 |
| CUD (%) | 26.7% (30) | 100% (13) | <0.0001 | 23.3% (30) | 100% (19) | <0.0001 |

The confusion matrix of DRAMA-BPD applied to MEX results is shown in Figure 3. As visible, 12 SA out of 13 are correctly identified, thus producing a single False Negative (FN) and a high Recall. On the other hand, 14 out of 19 NA are misclassified, causing a high number of False Positives (FP), and thus a low Precision. The classification resulted in the following global metrics: accuracy = 0.531, Precision = 0.462, Recall = 0.923, F1-score = 0.615 AUC = 0.593.

**Figure 3.**
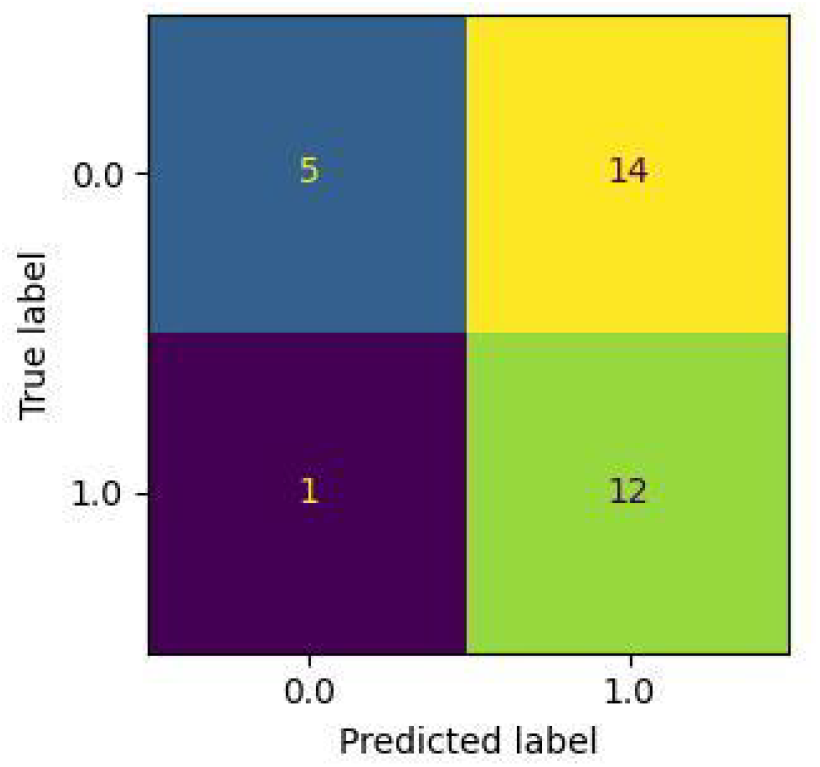
Confusion matrix of the DRAMA-BPD classification on the MEX dataset.

## 4. Discussion

### 4.1 Classifier pipeline

We performed a literature search to assess the impact of the important features MRI found by our algorithm.

#### Thickness of the right medial OFC

The OFC is involved in reward processing, pleasure, and happiness, and research evidence shows a decreased connectivity with hippocampal memory systems in depressed patients [31]. Our data reveal a significant difference between SA and NA BPD patients (p-value = 0.001), with a larger thickness for SA patients. Similar differences have been explored in the literature by Besteher et al. [32] and Ding et al. [33], but their results seem to disagree with our findings. It is however important to highlight some significant discrepancies, i.e., the different psychiatric pathology of the involved studies (BPD in our case, schizophrenia for [32], and MDD/BD for [33]), the different acquisition setups (3 T in our case, and 1.5 T in both the mentioned studies), and the different FS versions used to process MRI data.

#### FA average weight of AC

The AC is a bundle of nerve fibers in the brain that connects the two cerebral hemispheres, allowing them to coordinate various functions such as sensory perception, motor control, and cognitive processing [34]. Our data show a decreased FA for this region in SA patients, with a significant difference when compared with NA (p-value = 0.036). Literature does not show direct evidence to correlate the FA of the AC with suicidal behaviors in patients with psychiatric disorders.

#### Hypointensities of the WM

Hypointensities in T1-weighted images typically represent tissues with decreased signal intensity compared to surrounding ones. This can include various pathological features or anatomical structures, e.g., calcifications, hemorrhages, and scar tissues [35]. Our data present a significant difference between SA and NA BPD patients (p-value = 0.02), with a larger volume of WM hypointensities for SA patients. Grangeon et al. [36] performed a meta-analysis of MRI studies, concluding that patients with Deep WM Hyperintensities (DWMH) and PeriVentricular Hyperintensities (PVH) show a higher risk of suicidality, even though the underlying mechanisms are still to be uncovered. Our findings thus seem to agree with literature, highlighting the limitation of having hypointensities instead of hyperintensities.

#### Volume of the RC

The RC nucleus is involved in various functions related to movement, cognition, and emotion. Our findings indicate a lack of statistically significant difference for this region between SA and NA (p-value = 0.26), although a reduced volume is observed for SA. Research by Ho et al. [37] suggests that a smaller volume for RC is associated with greater implicit Suicidal Ideation (SI) in depressed adolescents. Wagner et al. [38] indicate that SA with a history of violent suicidal showed a decreased GM density in the right caudate. Although these studies employ different methodologies compared to ours, they converge on a common finding, suggesting that a reduced volume of the RC is associated with suicidal ideation.

#### Volume of the CCP

The corpus callosum (CC) plays a crucial role in interhemispheric communication, allowing for the transfer of information between the left and right cerebral hemispheres. Our data do not show a significant difference between SA and NA (p-value = 0.28), although a reduced volume is observed for SA. Gifuni et al. [25] investigated the relation between CC and suicidal vulnerability, revealing a significant reduced CC volume for BD, but no effects on suicidal history. Cyprien et al. [30] showed a reduced area size of the posterior third of the CC in subjects with a history of suicide, suggesting a possible role of CC in the pathophysiology of suicidal behavior. In light of the mentioned papers, our data is aligned with literature, and it is a further evidence that a reduced volume of CCP is related to mood disorders and/or suicidal behaviors.

#### Volume of the LCP

The left choroid plexus is a network of blood vessels surrounded by ependymal cells, which are specialized cells that produce cerebrospinal fluid (CSF). Our data show an increased volume for SA patients, but not a significant difference when compared with NA, with a p-value of 0.053. Literature shows no evidence of a relation between alteration in the volume of LCP and suicidality; however a link between alterations of this region and mood disorders seems to be present.

In summary of the presented results, it is important to note that the comparison with our work has limitations across all the mentioned articles. None of them focused on BPD individuals while analyzing suicidal behaviors. Additionally, only a few used a 3T equipment for MRI acquisition, and none of them employed our version of FS. Nevertheless, our findings align with literature for all volumetric features (i.e., hypointensities of the WM, volume of the RC, CCP, and LCP, 4 of the 6 MRI features), while a disagreement is observed for the sole thickness feature (thickness of medial OFC). It is challenging to draw conclusions about the FA average weight of the AC due to the lack of related papers. However, these findings suggest that the regions identified as important by the feature selection strategy are relevant for BPD subjects, thus validating our feature selection approach and strengthening the classifier results.

### 4.3 Independent dataset

Predictions on the MEX dataset show that out of the 13 SA, 12 are correctly identified. This results in 12 True Positives and a single False Negative (also known as a type 2 error), leading to a very high Recall of 0.923. Conversely, 14 of the 19 NA are mistakenly identified as SA, resulting in 5 True Negatives and 14 False Positives (also known as type 1 errors), thus with a very low Precision of 0.462. From a practical standpoint, these results indicate that the classifier correctly identifies most actual suicidal subjects, missing only a few (low type 2 errors). However, it also incorrectly labels a large number of non-suicidal subjects as suicidal (high type 1 errors). In summary, the classifier tends to overestimate the number of SA, sacrificing Precision for a high Recall. Generally speaking, high Recall is valuable in medical scenarios, because failing to detect positive instances could be dangerous. The downside is that incorrectly identified patients would undergo unnecessary follow-up tests or treatments, which can be costly and stressful. Efforts should be made to manage type 1 errors to avoid unnecessary procedures. It is also important to remember that DRAMA-BPD is intended as a second-opinion tool, thus clinicians could decide to apply further tests to discriminate between the two classes.

## 5. Limitations and future work

This work presents some limitations. The first is that the overall results, although in line with the literature, are not very high, reaching an F1-score of approximately 0.72 on the original dataset (CLM) and 0.62 on the independent one (MEX). While this is not a limitation per se, it shows that this instrument should still be considered a prototype and will require further development before being clinically reliable. The second limitation, related to the first one, is the small size of the CLM dataset. This is caused by several factors, such as the difficulties in identifying BPD individuals and the cost of MRI procedures, among others. The power analysis shows that both these limitations could be overcome by increasing the dataset size fourfold. This should provide a more powerful dataset and thus a more robust classifier, suitable for real-world usage. Another limitation is the comparison with the independent MEX dataset. All subjects in it are CUD individuals, while only a few subjects in the CLM dataset present drug addiction, and the majority of subjects in MEX are men, whereas CLM has approximately 85% women. Moreover, the feature numSPDC of the CLM dataset (number of lifetime admissions to SPDC) is not present in MEX as SPDCs do not exist in Mexico. Therefore, we used a similar variable, i.e., the P1 item of the ASI scale (“*How many times have you been treated for any psychological or emotional problem in a hospital?*”). This limitation could explain the relatively low performance. Future work would be related to the expansion of the CLM dataset and comparison with a more compatible independent dataset.

## 6. Conclusion

While suicide is a common cause of death among individuals with BPD, identifying subjects at risk remains a challenging task. In recent years, ML approaches for predicting suicide risk in patients with mental disorders have been developed. However, most of these approaches focus on different pathologies (e.g., MDD, psychosis, schizophrenia). The few that focus on BPD present some methodological limitations, such as overfitting and a lack of validation using external datasets. In this work, we aimed to overcome these limitations and developed DRAMA-BPD. By merging sociodemographic data, clinical information, emotional assessments, and MRI data, we built a dataset of 60 records with 276 features. We first applied a feature selection step, resulting in 7 features, 6 of which were derived from MRI data. We then implemented an SVM model and evaluated it using Grid Search with LOO cross-validation, resulting in the following metrics: Accuracy = 0.733, Precision = 0.750, Recall = 0.700, F1-score = 0.724, and AUC = 0.733. To validate the general applicability of DRAMA-BPD, we tested it on an independent dataset composed of 32 cocaine-addicted BPD subjects. The resulting metrics are: Accuracy = 0.531, Precision = 0.462, Recall = 0.923, F1-score = 0.615, and AUC = 0.593. While the overall AUC and F1-score are lower than the original metrics but still close to 0.6, it is important to note the significant imbalance between the high Recall (0.923) and low Precision (0.462). This demonstrates the potential of the proposed model but also indicates that further development and refinement are necessary to improve overall performance.

## Data Availability

All data produced in the present study are available upon reasonable request to the authors

